# Recent COVID-19 Vaccination Before Glioblastoma Surgery Is Associated With Longer Survival

**DOI:** 10.64898/2026.07.14.26358106

**Authors:** Sivakrishna C. Uppalapati, Danner W. Butler, Georges Bouobda, Elizabeth Liptrap, Philip G.R. Schmalz, Marshall T. Holland, Kristen Riley, Natalia Filippova, Louis B. Nabors, James M. Markert

**Author notes:** **Corresponding Author:** Sivakrishna C. Uppalapati, AB, Department of Neurosurgery, University of Alabama at Birmingham, 1702 2^nd^ Ave S, Birmingham, AL 35294.

## Abstract

**Background:** Glioblastoma remains resistant to most immune-based therapies. Surgery may create a perioperative window in which systemic immune activation and tumor antigen release intersect. We evaluated whether COVID-19 vaccination shortly before first glioblastoma surgery was associated with survival.

**Methods:** We performed a retrospective single-center cohort study of adults with newly diagnosed glioblastoma undergoing initial biopsy or resection from 2021 to 2025. The primary exposure was documented COVID-19 vaccination within 100 days before first tumor surgery. Overall survival was analyzed from surgery using Kaplan–Meier and Cox models, with 1:1 propensity matching and sensitivity analyses addressing treatment completion, calendar time, surgical selection, steroid exposure, immune-cell variables, COVID severity, and negative-control vaccination.

**Results:** The cohort included 187 patients: 64 perioperatively vaccinated and 123 non-perioperative comparators. Among vaccinated patients, 59/64 (92.2%) received mRNA vaccines; median vaccination-to-surgery interval was 81 days (IQR 71–90). Median overall survival was 743 days in vaccinated patients versus 318 days in comparators (unmatched HR 0.48, 95% CI 0.30–0.76; p=0.002). After 1:1 matching, median survival was 743 versus 349 days (HR 0.52, 95% CI 0.34–0.80). Sensitivity analyses accounting for adjuvant therapy, surgery year, extent of resection, steroid exposure, immune-cell measures, and COVID hospitalization were directionally consistent. Influenza vaccination was not associated with survival.

**Conclusions:** COVID-19 vaccination within 100 days before first glioblastoma surgery was associated with longer overall survival. These findings identify perioperative vaccination timing as a potentially relevant and modifiable variable in glioblastoma outcomes.

## Introduction

Glioblastoma (GBM) remains the most aggressive primary malignant brain tumor in adults despite maximal safe resection, radiation, temozolomide (TMZ), tumor-treating fields, and salvage systemic therapies.^1–4^ Durable responses to immunotherapy have been limited, in part because glioblastoma exists within a profoundly immunosuppressive tumor microenvironment characterized by tumor-associated macrophages and microglia, impaired antigen presentation, T-cell dysfunction, corticosteroid exposure, and systemic immune suppression.^5–8^ These features have made glioblastoma a paradigmatic “cold” tumor and have limited the effectiveness of checkpoint blockade, tumor vaccines, and other immune-based strategies.^5–8^

The operative environment, whether biopsy or resection, occupies a unique position in glioblastoma immunobiology.^9^ While resection is primarily performed for cytoreduction, diagnosis, and symptom control, it also creates a transient immunologic window in which tumor antigen release, wound healing, perioperative inflammation, corticosteroid exposure, lymphocyte dynamics, and adjuvant therapy initiation converge. This window may be particularly relevant for immune priming because antigen release from disrupted tumor tissue occurs while systemic innate and adaptive immune pathways are changing. Prior work has emphasized that surgical resection may provide a unique opportunity to break local immune tolerance and mount an antitumor immune response, although the perioperative immune environment remains incompletely understood.^9^

The COVID-19 vaccination era created an unplanned natural experiment in perioperative immune modulation. Unlike tumor-specific vaccines, SARS-CoV-2 vaccines were broadly administered, time-stamped, and frequently given near major clinical events, including cancer diagnosis and surgery. Importantly, mRNA vaccine platforms do more than generate antigen-specific humoral immunity: they can induce systemic innate immune activation, type I interferon signaling, dendritic cell activation, and downstream T-cell priming.^10,11^ Recent work has demonstrated that SARS-CoV-2 mRNA vaccines can sensitize tumors to immune checkpoint blockade in non-CNS cancers, with preclinical data showing increased type I interferon signaling and innate immune-cell priming of CD8-positive T cells targeting tumor-associated antigens.^12^ Whether such an immune-primed perioperative state has relevance in glioblastoma is unknown. Glioma patients can mount and maintain vaccine-induced immunity after COVID-19 vaccination, supporting the biologic plausibility that vaccine timing could influence immune tone even in this immunosuppressed population.^13^ However, to our knowledge, no study has evaluated whether COVID-19 vaccination shortly before initial glioblastoma surgery is associated with survival or perioperative outcomes. This question is clinically important because vaccination status is modifiable, routinely documented, and potentially actionable during preoperative counseling.

We performed a retrospective single-center cohort study of adults with newly diagnosed glioblastoma undergoing initial surgical treatment to determine whether perioperative COVID-19 vaccination within 100 days before first tumor surgery, defined as biopsy or resection, was associated with overall survival. We hypothesized that recent vaccination before surgery may reflect a clinically relevant immune-primed state, particularly in the setting of tumor antigen release and early adjuvant therapy planning. We further evaluated vaccine platform, perioperative length of stay and readmission, influenza vaccination as a negative control, and the robustness of the survival association after adjustment and propensity matching for clinical, tumor, frailty, performance status, steroid, COVID-severity, laboratory, and treatment-related variables.

## Methods

### Study design and cohort

We performed a retrospective single-center cohort study of adults with newly diagnosed glioblastoma treated at a large academic medical center between 2021 and 2025. Patients were included if they had a pathologic diagnosis of glioblastoma and underwent initial surgical management, defined as biopsy or resection, at our institution. The date of the first tumor operation was used as time zero for all survival analyses. Patients undergoing surgery for recurrent disease were excluded from the primary analysis to avoid mixing newly diagnosed and recurrence-era biology, treatment history, and survival risk. Clinical, operative, pathology, medication, laboratory, vaccination, and encounter-level data were obtained from institutional electronic health record extracts and supplemented by manual chart review when structured data were incomplete. Outside primary-care and pharmacy vaccination documentation was manually reviewed and incorporated when available. This study was reported in accordance with the Strengthening the Reporting of Observational Studies in Epidemiology (STROBE) guidelines for cohort studies.^14^

The University of Alabama at Birmingham Institutional Review Board reviewed this retrospective study and determined that it met the criteria for exempt human-subjects research. Under that determination, individual informed consent was not required.

### Exposure definition

The primary exposure was perioperative COVID-19 vaccination, defined as receipt of a documented COVID-19 vaccine within 100 days before first tumor surgery. Patients without a COVID-19 vaccine in this 100-day preoperative window were classified as non-perioperative comparators; this group included unvaccinated patients and patients vaccinated outside the prespecified perioperative window. Because vaccination status was defined using only vaccines administered before first tumor surgery, exposure status was assigned before time zero. Vaccines administered after surgery were not counted toward the primary exposure, thereby avoiding immortal-time bias. The interval from vaccination to surgery was calculated as the number of days from the most recent documented preoperative COVID-19 vaccine to first tumor surgery among patients with chart-confirmed vaccine dates. COVID-19 vaccine platform was categorized as mRNA or adenoviral-vector based on structured immunization records and chart-confirmed outside documentation. Because most perioperatively vaccinated patients received mRNA vaccines, mRNA-only analyses were performed as sensitivity analyses.

Influenza vaccination was evaluated as a negative-control exposure to test whether any observed association was specific to perioperative COVID vaccination rather than a nonspecific marker of vaccination behavior or healthcare engagement. Influenza vaccination was evaluated using both a one-year preoperative window and an analogous 100-day preoperative window when event counts permitted. Additional nonCOVID vaccine exposures, including pneumococcal, zoster, and any documented nonCOVID vaccine before surgery, were explored as additional negative-control proxies when available.

### Outcomes

The primary outcome was overall survival, defined as time from first tumor operation to death or last known follow-up. Vital status was determined from the institutional record and available follow-up documentation. Secondary perioperative outcomes included hospital length of stay after first tumor surgery and 30-day readmission. Readmission was defined as inpatient readmission within 30 days of discharge after first tumor surgery, based on encounter-level data and chart review when needed. Overall documented readmission was analyzed as an additional exploratory utilization outcome.

### Covariates

Covariates were selected a priori based on known glioblastoma prognostic factors, perioperative risk factors, and plausible confounders of vaccination status. Demographic and socioeconomic variables included age, sex, race/ethnicity when available, insurance/SES proxies, and Area Deprivation Index. Neighborhood socioeconomic deprivation was measured using the 2023 Area Deprivation Index national rank from the Neighborhood Atlas, linked to each patient’s available residential ZIP/ZIP+4 information in the electronic health record (EHR).^15,16^

Tumor and pathology variables included MGMT promoter methylation, IDH status, tumor location, multifocality, and deep/midline involvement.^2,17^ Surgical variables included biopsy versus resection as the primary surgical-treatment covariate. Among resected patients, extent of resection was abstracted when available and incorporated into secondary surgical-selection analyses using gross total resection (GTR) and non-gross-total resection categories. Tumor volume, eloquent-cortex involvement, TTFields adherence, exact salvage therapy sequencing, and exact temozolomide cycle counts were not uniformly available in structured data and were not included in the primary models. Perioperative clinical variables included mFI-5 frailty score, ASA class, note-derived Karnofsky Performance Status, ECOG performance status when available, steroid exposure, and COVID-19 infection/hospitalization severity.^18–21^ COVID infection and hospitalization before surgery were included as baseline covariates. Postoperative COVID events were analyzed descriptively and in sensitivity analyses but were not included as baseline confounders in the primary model.

Immune and hematologic variables included perioperative CBC with differential, including white blood cell count, absolute neutrophil count, absolute lymphocyte count, neutrophil-to-lymphocyte ratio, hemoglobin, and platelet count. KPS/ECOG values were extracted from clinic-note text and clinical event records. When multiple values were available, the closest available value to first tumor surgery was used for primary adjustment, and timing of KPS documentation was recorded to distinguish preoperative from peri/postoperative values in sensitivity analyses. Steroid exposure was derived from medication orders and perioperative medication documentation; dexamethasone was treated as the primary corticosteroid of interest, with other systemic steroids captured when present. Perioperative steroid exposure and CBC variables were defined using the closest available value or medication order within 30 days before through 30 days after first tumor surgery.

Adjuvant treatment variables were abstracted to evaluate treatment-completion bias. Radiotherapy variables included receipt of radiotherapy, available total dose/fractionation, and completion of standard-course radiotherapy when determinable. Temozolomide variables included receipt of temozolomide, medication-order documentation, and available markers of concurrent or adjuvant treatment exposure. Tumor-treating fields/Optune use was abstracted as a binary variable when documented. Because adjuvant radiotherapy, temozolomide, and tumor-treating fields occur after surgery and may lie on the causal pathway between perioperative immune state and survival, they were evaluated as secondary treatment-history and treatment-completion sensitivity variables rather than as primary baseline propensity-matching covariates.

### Statistical analysis

Baseline characteristics were summarized by perioperative vaccination status. Continuous variables were reported as mean ± standard deviation or median with interquartile range, depending on distribution. Categorical variables were reported as counts and percentages. Baseline balance after matching was evaluated using standardized mean differences, with an absolute SMD <0.10 considered well balanced.^22^

Unmatched overall survival was estimated using Kaplan–Meier methods and compared using Cox proportional hazards models.^23,24^ The primary unmatched analysis estimated the unadjusted association between perioperative COVID-19 vaccination and overall survival using Cox proportional hazards modeling. The main robustness analysis used 1:1 propensity-score matching to balance baseline variables including age, sex, socioeconomic variables, Area Deprivation Index, biopsy versus resection, mFI-5, MGMT, IDH, surgery year, time from diagnosis to surgery, ASA, KPS, and tumor anatomy.^25^ Matched survival was evaluated using Kaplan–Meier curves and Cox models in the matched cohort, with robust variance estimates used to account for matched pairs.

Because the exposure was defined strictly before surgery, the primary analysis was not modeled as a post-baseline or time-updated exposure. This was intentional: the biological question was whether a patient entered first surgery in a recently vaccinated immune state. Patients vaccinated after surgery did not contribute to the perioperative exposure group in the primary analysis.

### Sensitivity Analyses

Additional sensitivity analyses were performed to address potential residual confounding and plausible sources of bias. First, to evaluate treatment-completion bias, Cox models were repeated with treatment-history variables including radiotherapy receipt, temozolomide exposure, tumor-treating fields/Optune use, and a combined standard-of-care treatment proxy. A treatment-adjusted multivariable model incorporated age, MGMT status, KPS, extent-of-resection variables, surgery year, steroid exposure, radiotherapy, temozolomide, and tumor-treating fields use.

Second, to evaluate whether the 100-day exposure window was arbitrary, survival models were repeated using alternative preoperative COVID-19 vaccination windows of 30, 60, 90, 100, 180, and 365 days before first tumor surgery. Among perioperatively vaccinated patients with chart-confirmed dates, vaccine-to-surgery interval was additionally evaluated by timing strata and as a continuous variable scaled per 10-day interval. Third, to address calendar-time confounding during the COVID vaccination era, vaccination status was summarized by surgery year, and Cox models were repeated with surgery-year adjustment. Additional calendar-time sensitivity analyses included surgery-year stratified Cox models, within-year models when event counts permitted, and exact-year nearest-neighbor matched analyses.

Fourth, to address residual surgical-selection bias, analyses were repeated in the resection-only cohort and with extent-of-resection variables incorporated when available. Surgical sensitivity models incorporated biopsy versus resection, gross total resection/non-gross-total resection indicators, tumor location, multifocality, and deep/midline involvement. Fifth, to assess whether the association was explained by baseline immune status, steroid-related immunosuppression, or COVID disease severity, secondary models incorporated steroid exposure/dose, COVID infection/hospitalization severity, and perioperative CBC/immune-cell variables including ALC, ANC, NLR, WBC, platelet count, hemoglobin, and related differential measures. Additional models excluded patients with COVID hospitalization.

Sixth, landmark sensitivity analyses were performed at 30, 60, 90, and 180 days after first tumor surgery to assess whether early postoperative mortality disproportionately influenced the observed association. Patients who died before each landmark were excluded from the corresponding landmark analysis, and survival time was re-indexed from the landmark point. Seventh, negative-control analyses evaluated influenza vaccination within one year before surgery, influenza vaccination within 100 days before surgery, and additional non-COVID vaccine proxies when available. These analyses were intended to assess whether the perioperative COVID-19 vaccination association reflected a nonspecific healthy-vaccinee or healthcare-engagement effect.

Missingness was assessed for all covariates used in primary and sensitivity analyses. Variables with limited missingness were analyzed using complete-case methods in primary models, with missing/unknown categories used for molecular markers when clinically appropriate. A missingness table was generated for key demographic, molecular, clinical, treatment, vaccination, and outcome variables. Sensitivity analyses were performed to ensure that results were not driven by missing covariate data. The proportional hazards assumption was assessed using Schoenfeld residuals and log-minus-log survival plots.^26^ Two-sided p values were reported, with p<0.05 considered statistically significant.

### Exploratory computational shared-epitope analysis

After completion of the clinical analyses, we performed a post hoc, hypothesis-generating computational analysis to identify candidate peptide-level relationships between SARS-CoV-2 spike and GBM-associated antigens. These analyses did not inform cohort construction, exposure classification, clinical outcomes, covariate selection, or the primary survival models.

### Sequence-similarity and MHC-I binding analyses

A curated panel of 20 HLA class I-restricted GBM-associated or glioma-relevant peptides, together with a cytomegalovirus peptide control, was assembled from published GBM immunopeptidomic and vaccine studies.^30,31^ The full-length ancestral SARS-CoV-2 spike reference sequence (UniProt P0DTC2) was screened using a custom Python 3 workflow implemented with Biopython.^32^ For each candidate peptide, every same-length spike window was evaluated for exact amino-acid identity, BLOSUM62-positive substitutions, summed BLOSUM62 substitution score, and longest uninterrupted exact-match run.^33^ Candidates were ranked primarily by exact identity, followed by positive substitutions and BLOSUM62 score. A parallel exploratory screen of influenza A hemagglutinin sequences was used to identify viral comparator peptides. Two influenza HA-derived 9-mers, YNAELLVLL and DLWSYNAEL, were carried forward as comparator peptides for MHC-I and structural analyses.

Candidate GBM and viral peptides were evaluated using the Immune Epitope Database MHC-I binding-prediction resource with the IEDB-recommended NetMHCpan EL 4.1 method.^34,35^ Peptide lengths of 9 and 10 amino acids and a panel of commonly represented HLA-A and HLA-B alleles were evaluated. The primary mechanistic comparison focused on HLA-A*02:01 because the NLGN4X peptide NLDTLMTYV and SARS-CoV-2 spike peptide RLQSLQTYV were both predicted to bind this allele. NetMHCpan percentile rank was used as the principal measure, with rank <0.5 classified as strong predicted binding, rank 0.5–2.0 as weak-to-moderate predicted binding, and rank >2.0 as low-priority binding for the specified allele. These categories were used for candidate prioritization rather than formal hypothesis testing.

### Public-dataset validation of NLGN4X expression

NLGN4X expression was evaluated in two TCGA-GBM RNA-sequencing platforms and three independent Chinese Glioma Genome Atlas datasets.^36,37^ The TCGA datasets included the HiSeq RNA-sequencing cohort with available NLGN4X expression (n=152) and the Genome Analyzer RNA-sequencing cohort (n=525). CGGA analyses included primary WHO grade IV/GBM cases from mRNAseq_693 (n=140), mRNAseq_325 (n=85), and mRNA-array_301 (n=108). CGGA RNA-sequencing data were evaluated as log2(FPKM+1); TCGA values were analyzed in their downloaded log2 RSEM form; and the normalized microarray values were analyzed on their provided relative-expression scale. Because platforms used different normalization procedures, expression levels were summarized within datasets and were not pooled across platforms.

Spearman correlations were calculated between NLGN4X expression and predefined immune-expression signatures. The cytotoxic T-cell signature included CD8A, CD8B, GZMB, PRF1, and IFNG; the MHC-I antigen-presentation signature included HLA-A, HLA-B, HLA-C, B2M, TAP1, TAP2, PSMB8, and PSMB9; the interferon/CXCL signature included CXCL9, CXCL10, CXCL11, STAT1, ISG15, and MX1; and the checkpoint-inflammatory signature included CD274, PDCD1, LAG3, and HAVCR2. Signature scores were calculated from the mean available expression of component genes within each dataset. Individual gene-level Spearman correlations were also calculated. Benjamini-Hochberg correction was applied to correlation analyses. Exploratory univariable Cox models compared overall survival above versus below the dataset-specific median NLGN4X expression level; these analyses were intended to determine whether NLGN4X expression itself behaved as a reproducible prognostic marker rather than to establish a clinical expression threshold.

### Peptide-HLA structural modeling

HLA-A*02:01-bound structures were generated using the APE-Gen2.0 peptide-MHC class I modeling web server.^38^ Separate models were generated for NLGN4X NLDTLMTYV, SARS-CoV-2 spike RLQSLQTYV, influenza HA YNAELLVLL, and influenza HA DLWSYNAEL. Automatic anchor selection, primary/secondary anchor priority, anchor tolerance of 2, the default random-coordinate-descent step, Vinardo scoring, and rigid MHC settings were used. Energy minimization was not applied. The lowest-scoring conformation within each peptide-specific run was selected as the representative model; APE-Gen scores were used only for within-run conformation selection and were not interpreted as cross-peptide binding-affinity measurements.

Models were superimposed using HLA-A*02:01 heavy-chain Cα atoms. Peptide Cα root-mean-square deviation, peptide backbone root-mean-square deviation, position-specific side-chain centroid distances, and mean side-chain centroid distance across peptide positions P3–P8 were calculated using custom Python scripts. Shared residues at P2 and P9 were interpreted primarily as HLA anchor positions, whereas the middle and C-terminal peptide residues were examined as potential TCR-facing positions. Structural comparisons were descriptive; no inferential p values or prespecified RMSD threshold for molecular mimicry were used.

### TCR-footprint structural triage

To assess whether the modeled NLGN4X peptide was geometrically compatible with a known SARS-CoV-2-specific TCR footprint, we used the experimentally determined RLQ-specific TCR/HLA-A2/spike structure PDB 7N1E and the corresponding peptide-HLA reference PDB 7N1B.^39^ Each APE-Gen peptide-HLA model was aligned onto the HLA-A chain of 7N1E using heavy-chain Cα atoms from residues 1–180. The aligned peptide was then evaluated within the fixed experimental RLQ-specific TCR frame. Descriptive measures included peptide Cα and backbone RMSD relative to the experimental RLQSLQTYV peptide, mean P3–P8 side-chain centroid distance, peptide-TCR atom-pair contacts at ≤4.5 Å, and severe steric clashes at <2.5 Å. A same-method APE-Gen model of RLQSLQTYV served as an internal modeling control. This analysis assessed geometric compatibility with the experimental TCR footprint and was not interpreted as a prediction or demonstration of functional T-cell cross-reactivity. No molecular-dynamics simulation, free-energy calculation, or patient-specific HLA/TCR analysis was performed.

## Results

### Cohort and baseline characteristics

The primary cohort included 187 adults with newly diagnosed glioblastoma undergoing initial surgical treatment between 2021 and 2025, including 64 patients who received COVID-19 vaccination within 100 days before first tumor surgery and 123 non-perioperative comparator patients (Figure 1). Overall, 132 deaths occurred during follow-up. Deaths were less frequent in the perioperatively vaccinated group (26/64, 40.6%) than in the comparator group (106/123, 86.2%).

**Figure 1.**
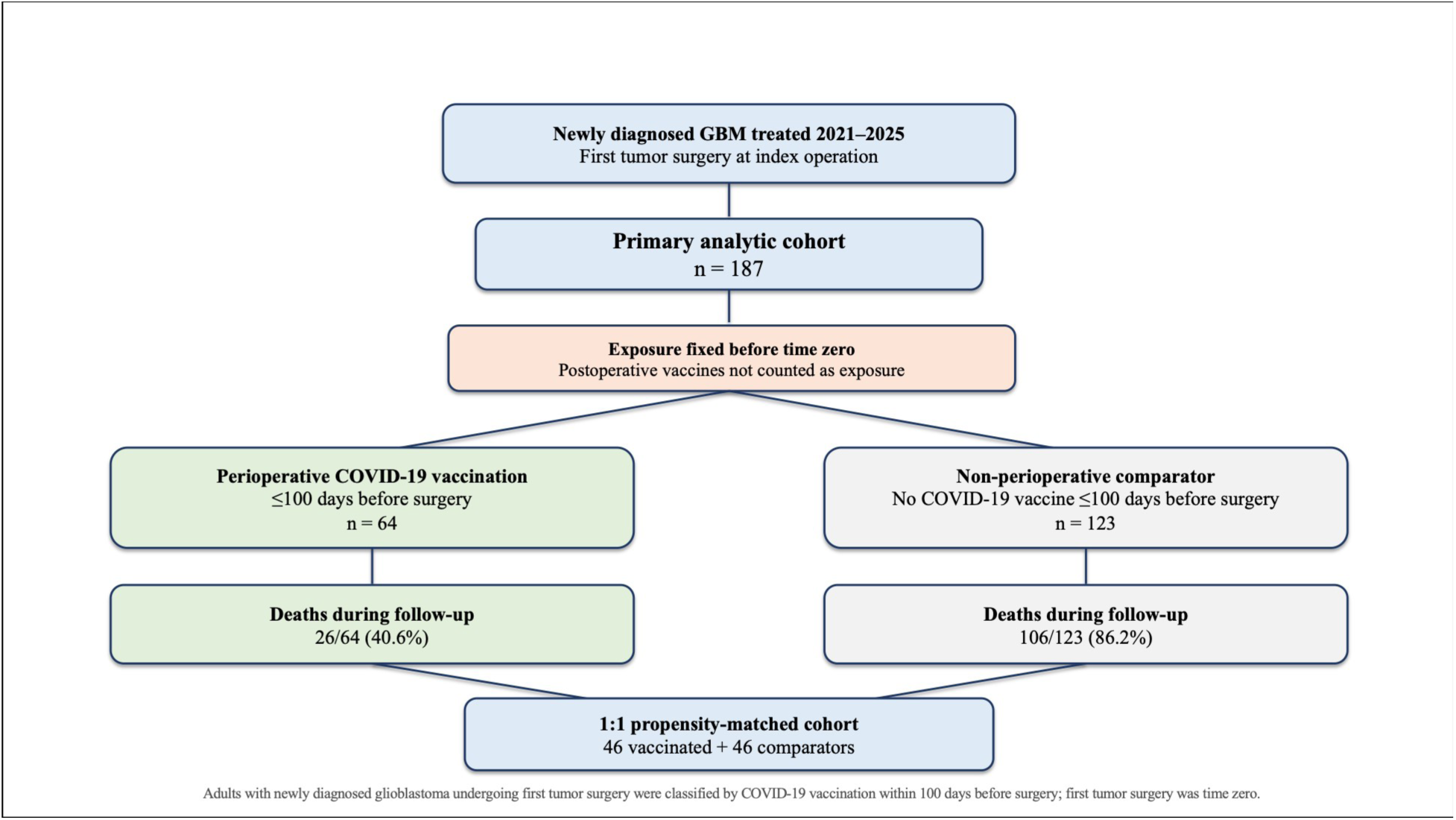
Primary Cohort (STROBE)

Before matching, perioperatively vaccinated patients were older on average (71.5 ± 10.0 vs 66.3 ± 9.2 years), while sex distribution was similar (male sex: 40/64 [62.5%] vs 74/123 [60.2%]). Surgical treatment was nearly identical between groups, with resection performed in 42/64 (65.6%) vaccinated patients and 81/123 (65.9%) comparator patients. MGMT methylation was also similar (23/64 [35.9%] vs 50/123 [40.7%]). Frailty, ASA, and performance status were comparable: median mFI-5 was 1 (IQR 1–2) in both groups, median ASA was 3 (IQR 3–3) in both groups, and median KPS was 80 (IQR 80–90) in vaccinated patients versus 80 (IQR 70–90) in comparators. Perioperative immune and steroid variables were also similar: median absolute lymphocyte count was 1.29 (IQR 0.88–1.83) versus 1.16 (IQR 0.80–1.72), maximum perioperative steroid dose was 5.5 mg daily (IQR 4.0–10.0) versus 8.0 mg daily (IQR 4.0–10.0), and COVID hospitalization was documented in 6/64 (9.4%) versus 10/123 (8.1%), respectively (Table 1).

**Table 1.** Primary Cohort Characteristics by Perioperative COVID-19 Vaccination Status.

| Characteristic | Perioperative COVID-19 vaccinated (n=64) | Non-perioperative comparator (n=123) |
| --- | --- | --- |
| **Demographic and surgical variables** |  |  |
| Age, years | 71.5 ± 10.0 | 66.3 ± 9.2 |
| Female sex | 24/64 (37.5%) | 49/123 (39.8%) |
| Male sex | 40/64 (62.5%) | 74/123 (60.2%) |
| Resection | 42/64 (65.6%) | 81/123 (65.9%) |
| Biopsy | 22/64 (34.4%) | 42/123 (34.1%) |
| MGMT methylated | 23/64 (35.9%) | 50/123 (40.7%) |
| mFI-5 | 1 (1–2) | 1 (1–2) |
| ASA class | 3 (3–3) | 3 (3–3) |
| KPS | 80 (80–90) | 80 (70–90) |
| **Perioperative immune/steroid/COVID variables** |  |  |
| Absolute lymphocyte count | 1.29 (0.88–1.83) | 1.16 (0.80–1.72) |
| Maximum perioperative steroid dose, mg daily | 5.5 (4.0–10.0) | 8.0 (4.0–10.0) |
| COVID hospitalization | 6/64 (9.4%) | 10/123 (8.1%) |
| **Adjuvant treatment proxies** |  |  |
| Radiotherapy receipt | 55/64 (85.9%) | 92/123 (74.8%) |
| Temozolomide exposure | 54/64 (84.4%) | 85/123 (69.1%) |
| Combined Stupp-regimen proxy | 52/64 (81.2%) | 82/123 (66.7%) |
| TTFields/Optune use | 6/64 (9.4%) | 12/123 (9.8%) |

*Values are mean ± SD, median (IQR), or n/N (%) unless otherwise indicated. Perioperative COVID-19 vaccination was defined as documented COVID-19 vaccination within 100 days before first tumor surgery. Treatment variables are postoperative treatment-history proxies and were evaluated in sensitivity analyses rather than used as primary baseline matching covariates.*

After 1:1 propensity matching, 46 perioperatively vaccinated patients were matched to 46 comparator patients. The matched cohort was well balanced across baseline covariates, including age, sex, resection status, frailty, MGMT, IDH, ASA, KPS, multifocality, deep/midline location, and ADI (Table 2). Absolute standardized mean differences were <0.10 for the key matched covariates, including sex (0.000), resection status (0.000), mFI-5 (0.043), MGMT methylation (0.000), IDH-wildtype status (0.000), ASA (0.031), KPS (0.001), multifocal tumor (0.082), deep/midline location (0.000), and ADI (0.014).

**Table 2.**
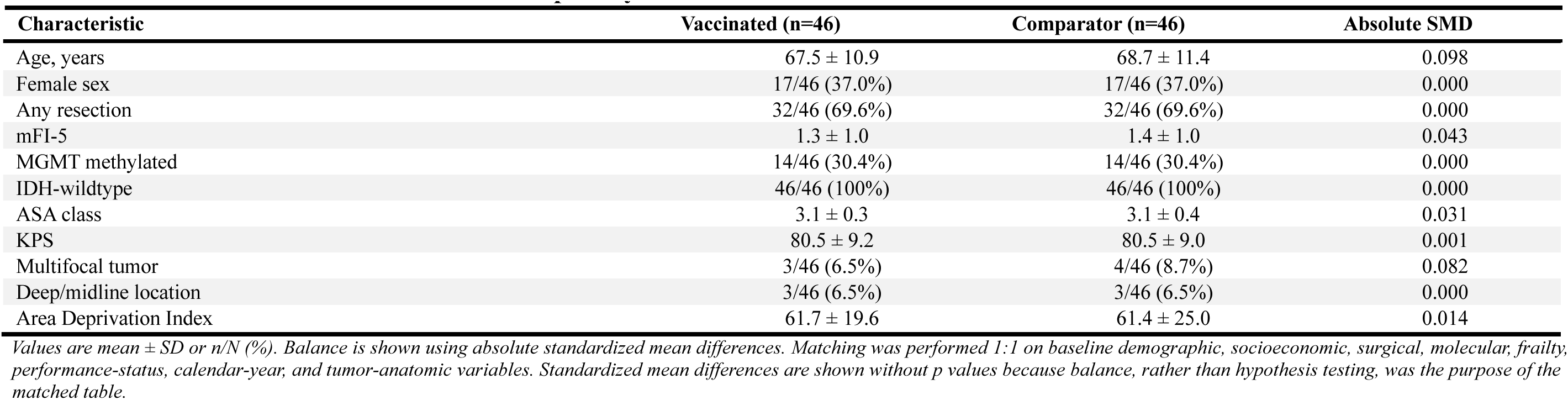
Baseline Characteristics of the 1:1 Propensity-Matched Cohort.

### Vaccine platform and timing

Among the 64 perioperatively vaccinated patients, the exposure was predominantly mRNA-based. Fifty-nine patients (92.2%) received an mRNA vaccine, whereas 5 patients (7.8%) received an adenoviral-vector vaccine (Figure 2). Median overall survival among adenoviral-vector recipients was 639 days, compared with 743 days among mRNA recipients. Given the very small adenoviral-vector subgroup (n=5), platform-specific survival differences were interpreted descriptively and were not used for formal inference. Chart-confirmed vaccine-to-surgery intervals were available for all 64 perioperatively vaccinated patients. The median interval from vaccination to first tumor surgery was 81 days (IQR 71–90, range 38–100).

**Figure 2 A+2B.**
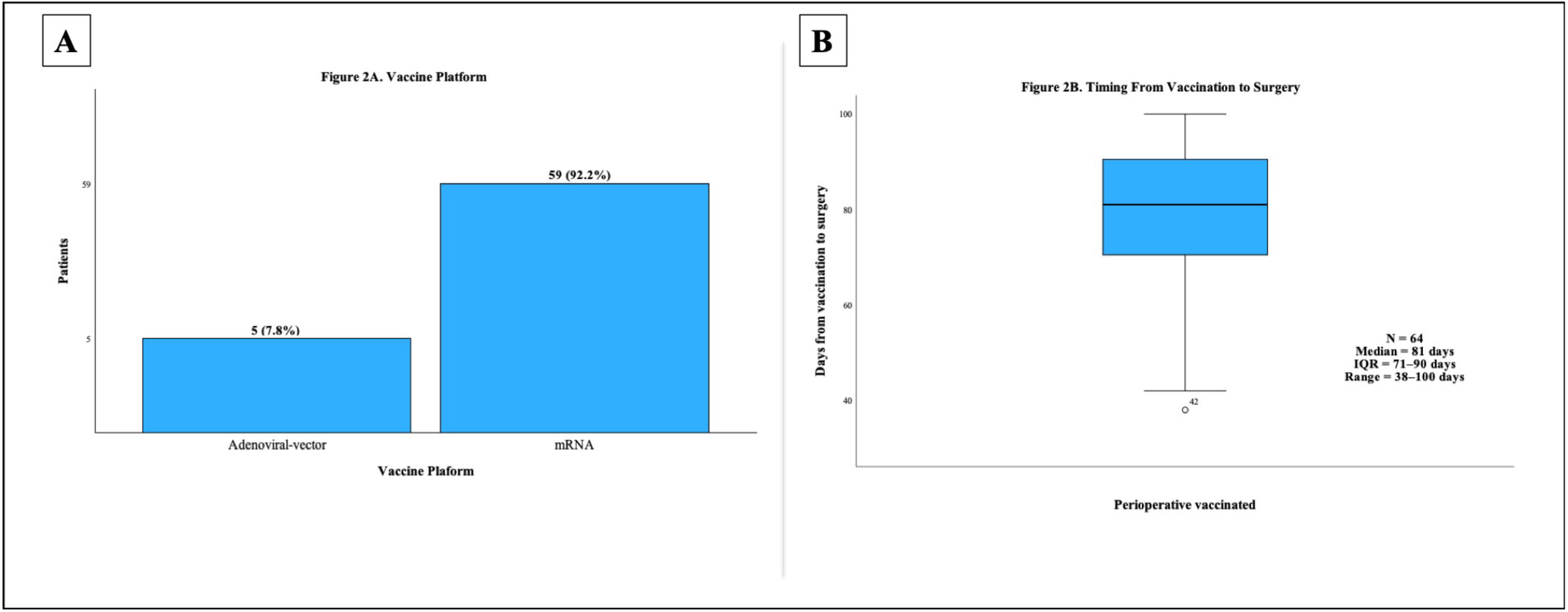
Perioperative Vaccinated Group

Alternative exposure-window analyses supported the 100-day perioperative definition. No patients were vaccinated within 30 days before surgery, so the 30-day window was not estimable. However, the association was already evident using narrower windows: 9 patients were vaccinated within 60 days before surgery (HR 0.39, 95% CI 0.19–0.81, p=0.012) and 48 patients were vaccinated within 90 days before surgery (HR 0.36, 95% CI 0.24–0.53, p=3.17×10⁻⁷). Using a 100-day window, 64 patients were exposed (HR 0.49, 95% CI 0.39–0.61, p=9.45×10⁻¹¹) (Figure 3). Among perioperatively vaccinated patients, vaccine-to-surgery interval was not associated with survival when modeled continuously (HR 0.99 per 10 days, p=0.966), and timing-bin analyses across the 38–100-day range showed directionally consistent estimates.

**Figure 3.**
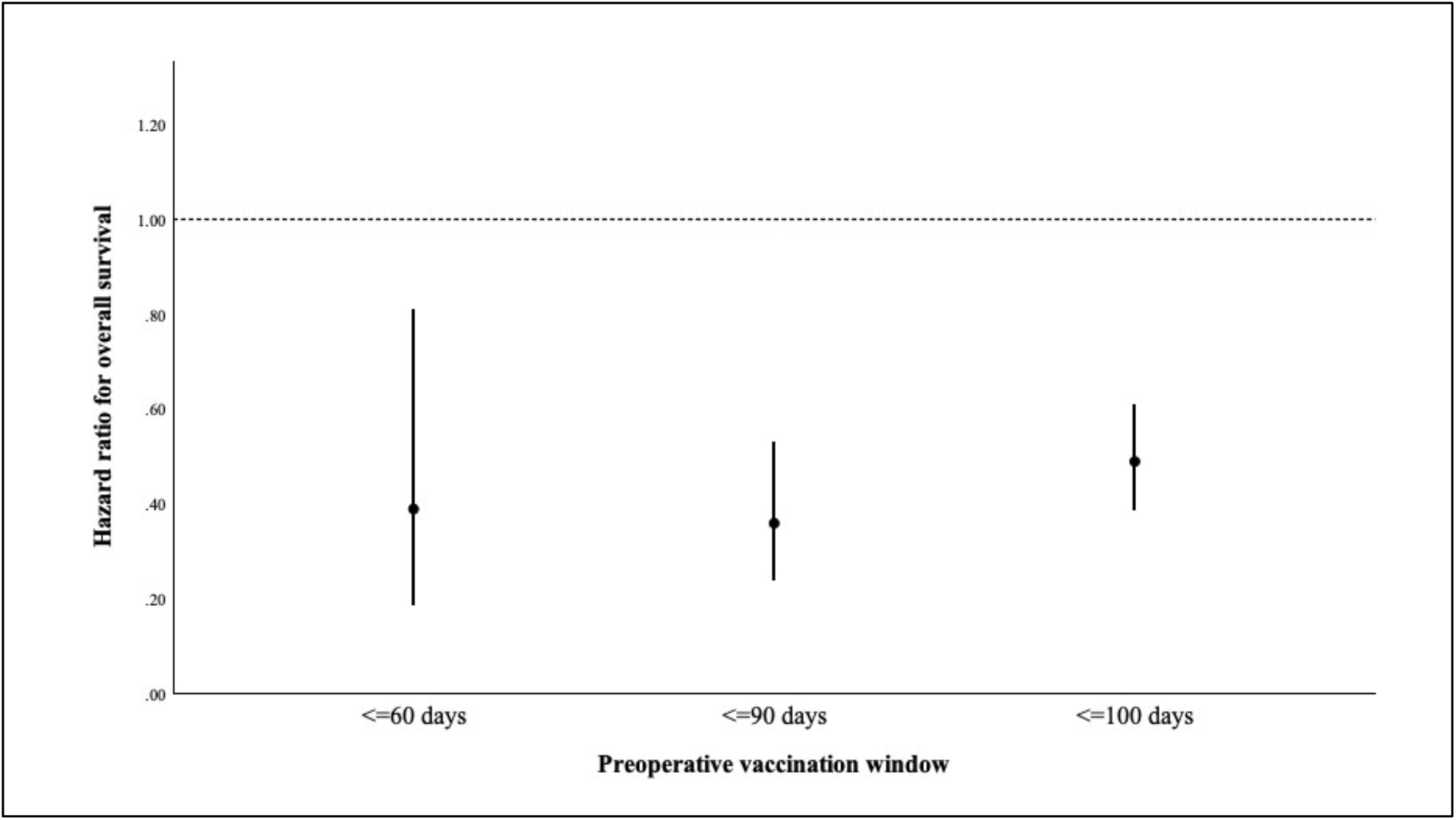
Exposure-window sensitivity

### Overall survival by perioperative COVID-19 vaccination status

In the unmatched cohort, median overall survival was 743 days in perioperatively vaccinated patients versus 318 days in non-perioperative comparators. Perioperative COVID-19 vaccination was associated with improved overall survival (unmatched HR 0.48, 95% CI 0.30–0.76, p=0.002; Figure 4A).

**Figure 4 A+B.**
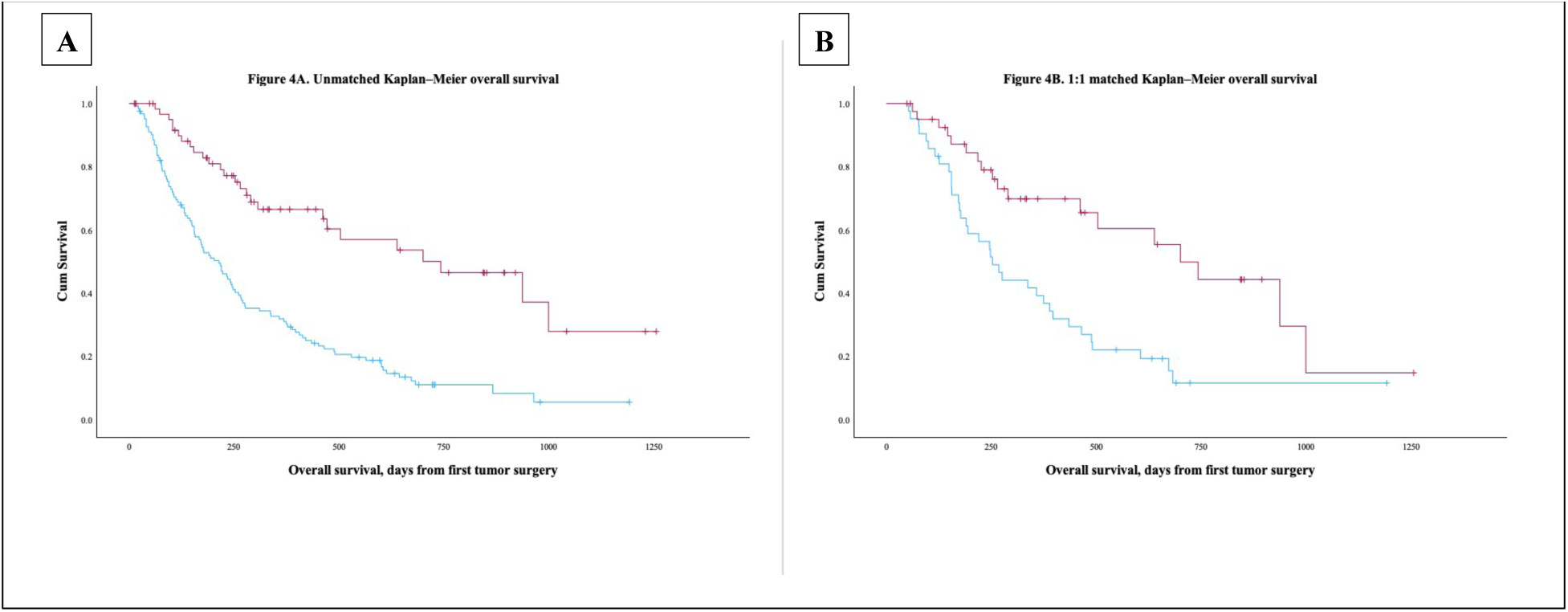
Unmatched and 1:1 matched overall survival

This survival difference was also observed after 1:1 propensity matching. In the matched cohort, median overall survival was 743 days among perioperatively vaccinated patients versus 349 days among matched comparators (HR 0.52, 95% CI 0.34–0.80, p<0.001; Figure 4B). The association was similar in the primary multivariable model adjusting for age, sex, biopsy versus resection, mFI-5, MGMT, and surgery year (HR 0.54, 95% CI 0.45–0.65, p=9.45×10⁻¹¹) and was not materially changed after additional adjustment for IDH, ASA, KPS, tumor anatomy, ADI, perioperative steroid exposure, ALC, and COVID hospitalization/severity (HR 0.53, 95% CI 0.41–0.68, p=5.36×10⁻⁷).

### Treatment-completion sensitivity analyses

Adjuvant treatment variables were evaluated to address treatment-completion bias. Perioperatively vaccinated patients were somewhat more likely to have documented standard-treatment proxies: radiotherapy receipt was documented in 55/64 (85.9%) vaccinated patients versus 92/123 (74.8%) comparators, temozolomide exposure in 54/64 (84.4%) versus 85/123 (69.1%), and a combined Stupp-regimen proxy in 52/64 (81.2%) versus 82/123 (66.7%). TTFields/Optune use was similar between groups (6/64 [9.4%] vs 12/123 [9.8%]).

Adjustment for treatment variables did not eliminate the survival association between perioperative COVID-19 vaccination and survival. In models incorporating treatment proxies individually, estimates remained favorable after adjustment for radiotherapy (HR 0.42, p=1.0×10⁻⁷), temozolomide exposure (HR 0.46, p=6.7×10⁻⁶), TTFields/Optune use (HR 0.49, p=5.9×10⁻⁷), and the combined Stupp-regimen proxy (HR 0.46, p=2.8×10⁻⁵). In a comprehensive treatment-adjusted multivariable model including age, MGMT, KPS, extent-of-resection variables, surgery year, steroid exposure, radiotherapy, temozolomide, and TTFields/Optune use, perioperative vaccination remained associated with improved survival (HR 0.51, 95% CI 0.39–0.67, p=1.02×10⁻⁶). With additional ASA, ADI, and tumor-anatomy adjustment, the estimate remained similar (HR 0.50, 95% CI 0.37–0.67, p=1.11×10⁻⁵) (Table 3; Figure 5).

**Figure 5.**
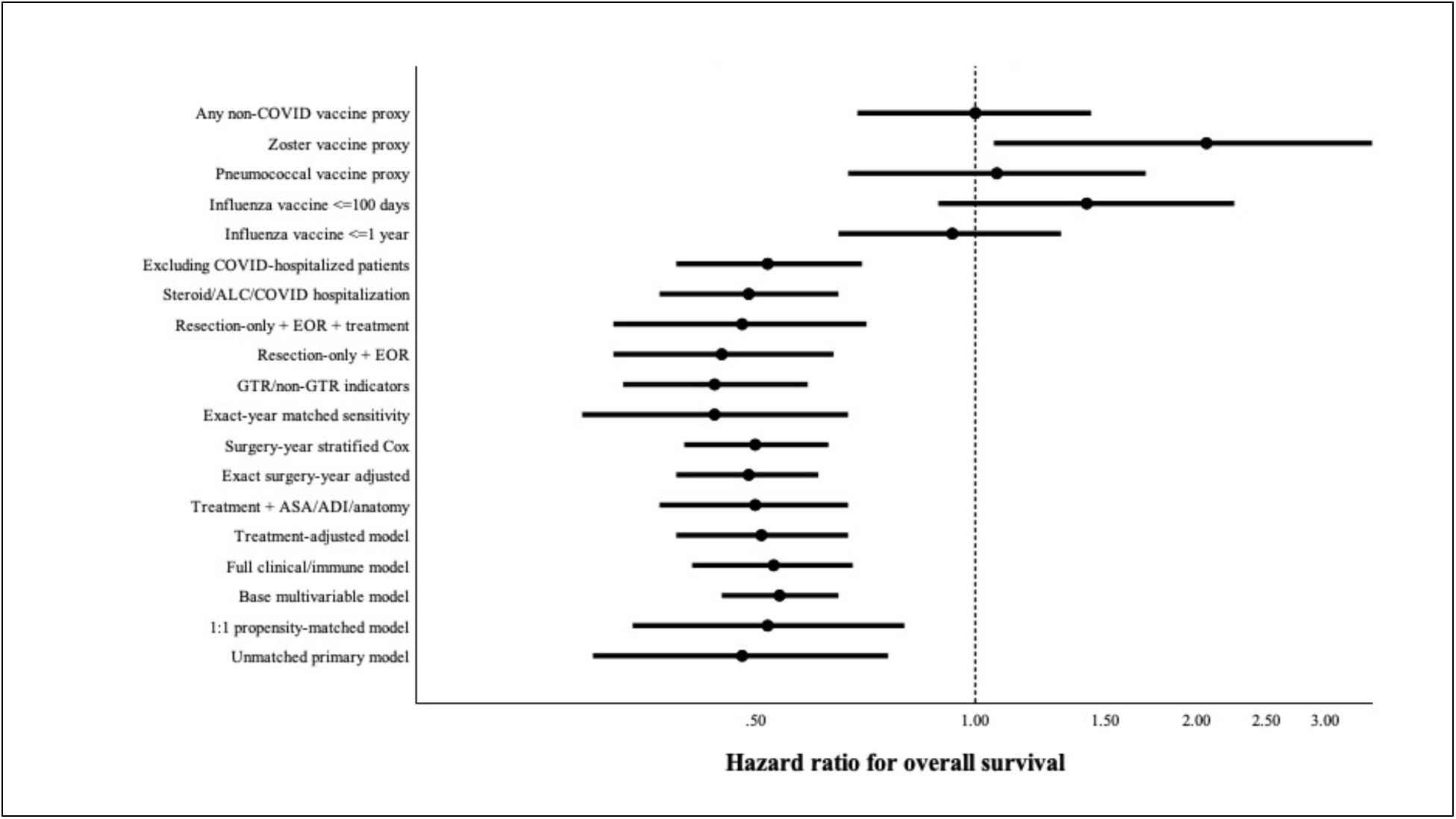
Robustness and negative-control sensitivity analyses

**Table 3.** Survival and Sensitivity Analyses for Perioperative COVID-19 Vaccination.

| Analysis | Cohort / model | HR | 95% CI | P value |
| --- | --- | --- | --- | --- |
| **Primary survival analyses** |  |  |  |  |
| Unmatched primary model | Full cohort | 0.48 | 0.30–0.76 | 0.002 |
| 1:1 propensity-matched model | Matched cohort | 0.52 | 0.34–0.80 | <0.001 |
| Base multivariable model | Age, sex, biopsy/resection, mFI-5, MGMT, surgery year | 0.54 | 0.45–0.65 | $9.45 \times 10^{-11}$ |
| Full clinical/immune model | Additional IDH, ASA, KPS, tumor anatomy, ADI, steroid exposure, ALC, COVID hospitalization/severity | 0.53 | 0.41–0.68 | $5.36 \times 10^{-7}$ |
| **Exposure-window analyses** |  |  |  |  |
| COVID vaccine $\leq 60$ days before surgery | 9 exposed | 0.39 | 0.19–0.81 | 0.012 |
| COVID vaccine $\leq 90$ days before surgery | 48 exposed | 0.36 | 0.24–0.53 | $3.17 \times 10^{-7}$ |
| COVID vaccine $\leq 100$ days before surgery | 64 exposed | 0.49 | 0.39–0.61 | $9.45 \times 10^{-11}$ |
| **Treatment-completion sensitivity analyses** |  |  |  |  |
| Treatment-adjusted model | Age, MGMT, KPS, EOR variables, surgery year, steroid exposure, RT, TMZ, TTFields | 0.51 | 0.39–0.67 | $1.02 \times 10^{-6}$ |
| Treatment + ASA/ADI/anatomy model | Treatment-adjusted model plus ASA, ADI, tumor anatomy | 0.50 | 0.37–0.67 | $1.11 \times 10^{-5}$ |
| **Calendar-time sensitivity analyses** |  |  |  |  |
| Exact surgery-year adjusted | Surgery-year adjustment | 0.49 | 0.39–0.61 | $3.11 \times 10^{-11}$ |
| Surgery-year stratified Cox model | Stratified by surgery year | 0.50 | 0.40–0.63 | $2.4 \times 10^{-9}$ |
| Exact-year nearest-neighbor matched sensitivity | Exact-year matched cohort | 0.44 | 0.29–0.67 | 0.00053 |
| **Surgical-selection sensitivity analyses** |  |  |  |  |
| GTR/non-GTR indicators | Model incorporating EOR indicators | 0.44 | 0.33–0.59 | $3.46 \times 10^{-8}$ |
| Resection-only + EOR | Resection-only cohort | 0.45 | 0.32–0.64 | $1.71 \times 10^{-5}$ |
| Resection-only + EOR + treatment | Resection-only cohort with treatment proxies | 0.48 | 0.32–0.71 | 0.00023 |
| **Immune, steroid, and COVID-severity sensitivity analyses** |  |  |  |  |
| Steroid/ALC/COVID hospitalization model | Adjusted for steroid exposure, ALC, COVID hospitalization | 0.49 | 0.37–0.65 | $5.36 \times 10^{-7}$ |
| Excluding COVID-hospitalized patients | COVID hospitalization excluded | 0.52 | 0.39–0.70 | $8.53 \times 10^{-6}$ |
| **Negative-control vaccination analyses** |  |  |  |  |
| Influenza vaccination $\leq 1$ year before surgery | Negative control | 0.93 | 0.65–1.31 | 0.664 |
| Influenza vaccination $\leq 100$ days before surgery | Negative control | 1.42 | 0.89–2.26 | 0.143 |
| Pneumococcal vaccine proxy | Negative-control proxy | 1.07 | 0.67–1.71 | 0.784 |
| Zoster vaccine proxy | Negative-control proxy | 2.07 | 1.06–4.03 | 0.034 |
| Any non-COVID vaccine proxy | Negative-control proxy | 1.00 | 0.69–1.44 | 0.981 |
| **Landmark analyses** |  |  |  |  |
| 30-day landmark | Excluding deaths before 30 days | 0.51 | 0.41–0.63 | $2.77 \times 10^{-10}$ |
| 60-day landmark | Excluding deaths before 60 days | 0.54 | 0.44–0.67 | $1.42 \times 10^{-8}$ |
| 90-day landmark | Excluding deaths before 90 days | 0.55 | 0.44–0.69 | $1.23 \times 10^{-7}$ |
| 180-day landmark | Excluding deaths before 180 days | 0.54 | 0.40–0.73 | $5.91 \times 10^{-5}$ |

*Hazard ratios below 1.0 favor perioperative COVID-19 vaccination. The primary exposure was COVID-19 vaccination within 100 days before first tumor surgery. Treatment variables were evaluated as secondary treatment-history sensitivity variables because they occur after surgery and may partially mediate survival. EOR indicates extent of resection; RT, radiotherapy; TMZ, temozolomide; ALC, absolute lymphocyte count; ADI, Area Deprivation Index.*

### Calendar-time sensitivity analyses

Because the cohort spanned the COVID vaccination era, surgery year was evaluated as a potential confounder. Perioperative vaccination was represented across all surgery years: in 2022, 24 vaccinated patients and 25 comparators were included; in 2023, 15 vaccinated patients and 45 comparators; in 2024, 10 vaccinated patients and 40 comparators; and in 2025, 15 vaccinated patients and 13 comparators (Figure 6).

**Figure 6.**
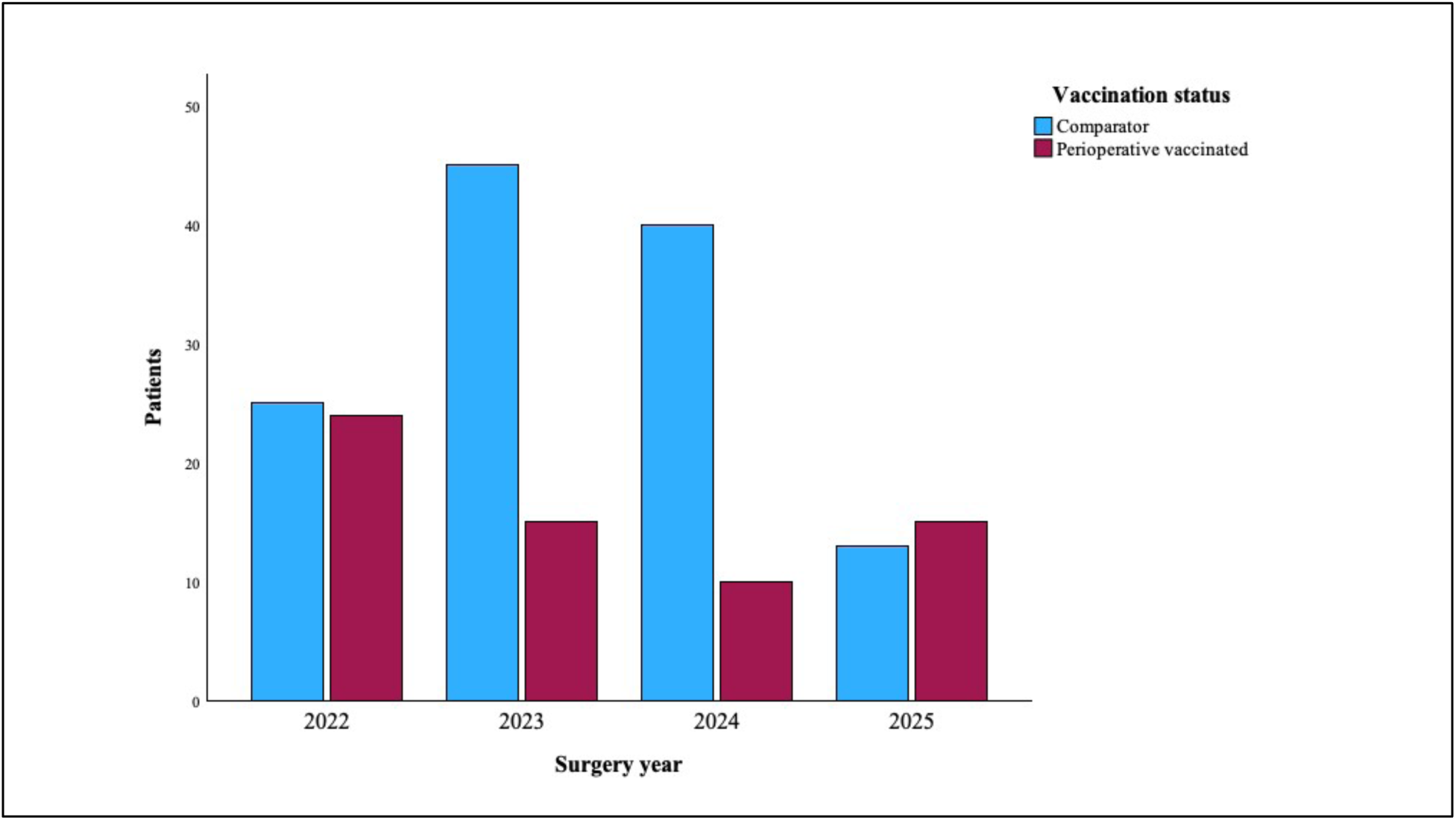
Calendar-time distribution

Calendar-time adjustment for exact surgery year yielded similar results (HR 0.49, 95% CI 0.39–0.61, p=3.11×10⁻¹¹) and in a surgery-year–stratified Cox model (HR 0.50, 95% CI 0.40–0.63, p=2.4×10⁻⁹). Within-year estimates remained directionally favorable across each year with sufficient events: 2022 HR 0.46 (p=0.0097), 2023 HR 0.50 (p=6.85×10⁻⁵), 2024 HR 0.33 (p=0.048), and 2025 HR 0.36 (p=0.049). In an exact-year nearest-neighbor matched sensitivity analysis, perioperative vaccination remained associated with improved survival (HR 0.44, 95% CI 0.29–0.67, p=0.00053).

### Surgical-selection and extent-of-resection sensitivity analyses

Surgical treatment was balanced in the primary cohort and included in the propensity-matching model. Additional surgical-selection analyses were performed to evaluate whether the association was driven by extent of surgery. In models incorporating gross-total-resection and non-gross-total-resection indicators, perioperative vaccination remained associated with improved survival (HR 0.44, 95% CI 0.33–0.59, p=3.46×10⁻⁸). In the resection-only cohort, the association remained after incorporating extent-of-resection proxies (HR 0.45, 95% CI 0.32–0.64, p=1.71×10⁻⁵) and after additional adjustment for treatment proxies (HR 0.48, 95% CI 0.32–0.71, p=0.00023).

### Immune, steroid, and COVID-severity sensitivity analyses

Secondary models incorporated perioperative steroid exposure, ALC, and COVID hospitalization/severity to evaluate whether the survival association was explained by baseline immune status, steroid-related immunosuppression, or COVID disease severity. Adjustment for steroid exposure, ALC, and COVID hospitalization did not materially alter the estimate (HR 0.49, 95% CI 0.37–0.65, p=5.36×10⁻⁷). The association also persisted after excluding patients with COVID hospitalization (HR 0.52, 95% CI 0.39–0.70, p=8.53×10⁻⁶).

### Negative-control vaccination analyses

Influenza vaccination within one year before surgery was not associated with improved overall survival (HR 0.93, 95% CI 0.65–1.31, p=0.664). Using an analogous 100-day preoperative window, influenza vaccination also was not associated with improved survival (HR 1.42, 95% CI 0.89–2.26, p=0.143). Additional non-COVID vaccine proxies likewise did not reproduce the COVID-vaccination association: pneumococcal vaccination was not associated with survival (HR 1.07, 95% CI 0.67–1.71, p=0.784), zoster vaccination was associated with worse rather than improved survival (HR 2.07, 95% CI 1.06–4.03, p=0.034), and any documented non-COVID vaccine before surgery showed no association (HR 1.00, 95% CI 0.69–1.44, p=0.981). These negative-control analyses did not support a generalized “vaccinated patients live longer” pattern.

### Landmark analyses

Landmark analyses were performed to assess whether early postoperative mortality disproportionately drove the observed association. The association remained significant after excluding patients who died before 30 days (HR 0.51, 95% CI 0.41–0.63, p=2.77×10⁻¹⁰), 60 days (HR 0.54, 95% CI 0.44–0.67, p=1.42×10⁻⁸), 90 days (HR 0.55, 95% CI 0.44–0.69, p=1.23×10⁻⁷), and 180 days (HR 0.54, 95% CI 0.40–0.73, p=5.91×10⁻⁵).

### Perioperative utilization

In the overall cohort, perioperatively vaccinated patients had shorter postoperative hospitalization. Median length of stay was 4 days in the vaccinated group compared with 5 days in comparators (p=0.008; Figure 7A). This difference was observed despite nearly identical biopsy-versus-resection distribution between groups, and the LOS distribution showed fewer prolonged postoperative stays among vaccinated patients. Thirty-day readmission was numerically lower in vaccinated patients (4/64 [6.3%] vs 15/123 [12.2%]), although this difference was not statistically significant (OR 0.48, 95% CI 0.15–1.51, p=0.307; Figure 7B). Among the 19 total 30-day readmissions, categories included neurologic decline, edema, hemorrhage, or focal neurologic symptoms (n=5), tumor-related or other neurologic deterioration (n=7), wound/SSI/CSF-related issues (n=3), seizure (n=2), and systemic infection, encephalopathy, or medical deterioration (n=2). Overall documented readmission was also numerically lower in the vaccinated group (6.3% vs 13.8%), supporting a consistent but secondary perioperative-utilization trend.

**Figure 7 A+B.**
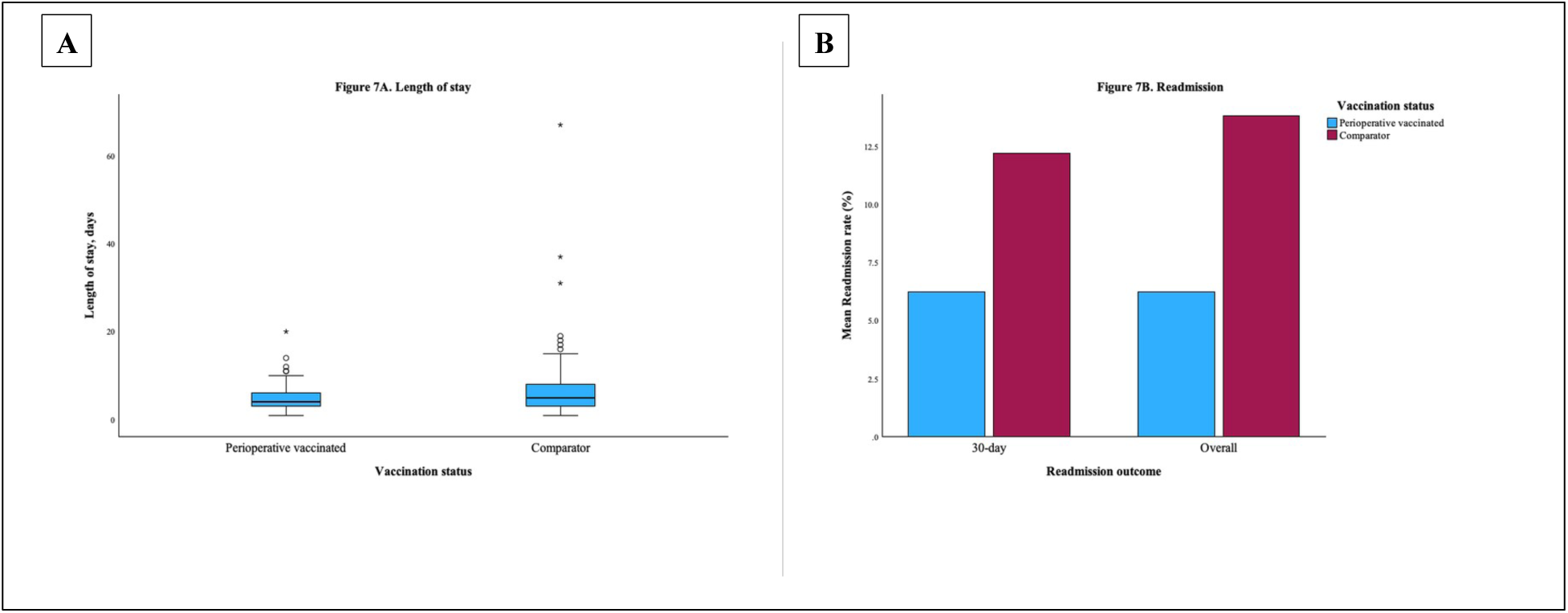
Length of stay and Readmission

### Exploratory shared-epitope, expression, and structural analyses

The sequence-similarity screen identified several partial matches between SARS-CoV-2 spike and the curated GBM-associated peptide panel. The cleanest same-HLA candidate was the NLGN4X peptide NLDTLMTYV and the SARS-CoV-2 spike peptide RLQSLQTYV, corresponding to spike residues 1000–1008. The peptides shared 5 of 9 amino acids at P2, P5, P7, P8, and P9. This included the shared HLA-A*02:01-compatible anchor architecture at P2 leucine and P9 valine, together with an identical C-terminal T-Y-V motif at P7–P9.

Both peptides were strong predicted HLA-A*02:01 ligands. NetMHCpan EL 4.1 percentile ranks were 0.09 for NLGN4X NLDTLMTYV and 0.05 for spike RLQSLQTYV. In the tested influenza comparator set, HA YNAELLVLL had weaker predicted HLA-A*02:01 presentation (percentile rank 2.70), whereas HA DLWSYNAEL had weak-to-moderate predicted binding (percentile rank 0.95). A secondary IL13Rα2/spike-region candidate was less clean in the same HLA context: IL13Rα2 WLPFGFILI had HLA-A*02:01 percentile rank 1.40, the exact spike 9-mer WLGFIAGLI had rank 9.60, and the overlapping spike 10-mer YIWLGFIAGL had rank 1.30.

NLGN4X expression was detected across all evaluated TCGA and CGGA GBM datasets. In CGGA mRNAseq_693 primary GBM cases, median NLGN4X expression was 2.70 log2(FPKM+1), and 130/140 tumors (92.9%) had raw FPKM ≥1. In CGGA mRNAseq_325, median expression was 3.46 log2(FPKM+1), and all 85 tumors had raw FPKM ≥1. NLGN4X also had nonzero normalized expression in all analyzed CGGA microarray cases and both TCGA-GBM expression cohorts. These results supported tumor-level expression of the source gene but did not establish presentation of the NLDTLMTYV peptide on individual tumors.

NLGN4X-high tumors did not show a consistent immune-inflamed phenotype in bulk transcriptomic data. Correlations between NLGN4X and the cytotoxic T-cell signature were inverse in all five datasets, with Spearman ρ values ranging from −0.22 to −0.44. Associations with MHC-I antigen-presentation, interferon/CXCL, and checkpoint-inflammatory signatures were heterogeneous across platforms. NLGN4X expression alone was not significantly associated with overall survival in median-split analyses of any TCGA or CGGA cohort; hazard ratios ranged from 0.80 to 1.45, with all p values >0.07.

APE-Gen modeling showed close structural concordance between HLA-A*02:01-bound NLGN4X NLDTLMTYV and spike RLQSLQTYV. After HLA heavy-chain alignment, peptide backbone RMSD was 0.33 Å, peptide Cα RMSD was 0.34 Å, and the mean P3–P8 side-chain centroid distance was 0.83 Å. The shared P5 leucine, P7 threonine, and P8 tyrosine side-chain centroid distances were 0.69 Å, 0.30 Å, and 0.60 Å, respectively. The tested influenza HA comparators showed less structural concordance with NLGN4X: backbone RMSD was 0.95 Å for YNAELLVLL and 1.10 Å for DLWSYNAEL, with mean P3–P8 side-chain distances of 2.22 Å and 2.44 Å, respectively.

When placed into the experimental RLQ-specific TCR/HLA-A2 frame from PDB 7N1E, the modeled NLGN4X peptide retained close geometric similarity to the experimental spike peptide. NLGN4X had a peptide backbone RMSD of 0.51 Å relative to experimental RLQSLQTYV, a mean P3–P8 side-chain centroid distance of 0.91 Å, and two atom-pair clashes at <2.5 Å. The same-method APE-Gen spike control had backbone RMSD of 0.45 Å, mean P3–P8 side-chain distance of 0.83 Å, and three clashes at <2.5 Å. The influenza YNAELLVLL and DLWSYNAEL models were less compatible in this geometric comparison, with backbone RMSD values of 0.91 Å and 1.35 Å and 6 and 29 severe clashes, respectively. The principal sequence uncertainty within the known RLQ-specific TCR footprint was P6, where spike glutamine was replaced by methionine in NLGN4X. Collectively, these findings supported a plausible shared HLA-A*02:01/TCR-facing geometry but did not demonstrate functional cross-recognition.

### Missingness and data completeness

Missingness was summarized for all variables used in primary and sensitivity analyses (Supplementary Table 1). Chart-confirmed vaccine-to-surgery intervals were available for all 64/64 perioperatively vaccinated patients in the final analysis. IDH status was missing or unknown in 1/187 (0.5%) patients. Missingness was greater for some treatment-intensity variables, including exact temozolomide cycle count, radiotherapy interruption/completion details, and TTFields adherence; therefore, treatment analyses used documented receipt/completion proxies, and incompletely captured treatment-intensity measures were treated as sensitivity variables rather than primary matching covariates.

## Discussion

### Perioperative vaccination and overall survival

In this retrospective cohort of adults with newly diagnosed glioblastoma, COVID-19 vaccination within 100 days before first tumor surgery was associated with improved overall survival. The association remained present in the unmatched cohort and continued to exist after 1:1 propensity matching across demographic, socioeconomic, surgical, molecular, frailty, performance-status, and tumor-anatomic variables. The exposure was predominantly mRNA-based, and influenza vaccination was not associated with improved survival. Perioperatively vaccinated patients also had shorter postoperative length of stay and numerically lower 30-day readmission.

The central finding is not simply that vaccinated patients lived longer. The more specific observation is that recent vaccination before first GBM surgery was associated with a survival advantage that was directionally consistent across analyses addressing treatment completion, calendar time, surgical selection, steroid exposure, immune-cell variables, COVID severity, and negative-control vaccination. This is important because GBM is profoundly immunosuppressive, and most immune-based strategies have failed to produce durable benefit in unselected patients.^7,8,29^ The present data suggest that the timing of systemic immune activation relative to surgery may be an underrecognized variable in GBM outcomes.

### Treatment Completion

A major concern is that perioperatively vaccinated patients may have been more likely to complete standard GBM therapy. This is a plausible source of bias because receipt of radiotherapy, temozolomide, and tumor-treating fields can strongly influence survival. In our cohort, perioperatively vaccinated patients did have somewhat higher documented rates of radiotherapy receipt, temozolomide exposure, and a combined Stupp-regimen proxy, although TTFields/Optune use was similar between groups.

Importantly, the survival association persisted after accounting for these treatment variables. In treatment-adjusted models, perioperative vaccination remained associated with improved survival after incorporating radiotherapy receipt, temozolomide exposure, TTFields/Optune use, and a combined Stupp-regimen proxy. In a multivariable model incorporating age, MGMT, KPS, extent-of-resection variables, surgery year, steroid exposure, radiotherapy, temozolomide, and TTFields/Optune use, the association remained significant. These analyses do not eliminate all treatment-related confounding, particularly because exact temozolomide cycle counts, radiotherapy interruptions, and TTFields adherence were not uniformly available. However, they argue against the interpretation that the observed survival difference is explained solely by differential receipt of standard adjuvant therapy.

### Vaccine timing

The 100-day exposure window was selected to capture recent vaccination before first tumor surgery rather than remote vaccination history. Sensitivity analyses supported this perioperative framing. Although no patients were vaccinated within 30 days before surgery, the association was already present among patients vaccinated within 60 days and remained strong within 90 days. The prespecified 100-day window included all 64 perioperatively vaccinated patients, with a median vaccine-to-surgery interval of 81 days.

Within the perioperatively vaccinated group, vaccine-to-surgery interval was not associated with survival when modeled continuously, and timing-bin analyses across the 38–100 day range showed directionally consistent estimates. These findings reduce concern that the association was driven by a narrow, data-selected timing pocket. Rather, the data support a broader perioperative window in which recent vaccination before surgery may be clinically relevant.

### Calendar time

Calendar-time confounding is a major concern in a cohort spanning the COVID vaccination era. Vaccine availability, variant waves, healthcare utilization, molecular testing, supportive care, and referral patterns all changed between 2021 and 2025. In our cohort, perioperative vaccination was represented across multiple surgery years rather than being confined to a single calendar period.

Results were similar after exact surgery-year adjustment, surgery-year stratification, and exact-year matched sensitivity analysis. Within-year estimates remained directionally favorable across each year with sufficient events. These findings do not completely eliminate secular-trend bias, but they substantially reduce the likelihood that the survival association is simply an artifact of later-era care or improved supportive treatment over time.

### Surgical selection

Surgical selection bias is another central concern in observational GBM survival studies. Patients selected for resection often have more favorable tumor anatomy, better functional status, and longer expected survival than patients selected for biopsy.^2^ If perioperatively vaccinated patients were more likely to undergo resection, the observed association could be an artifact.

That does not appear to explain the present finding. The proportion undergoing resection rather than biopsy was essentially identical between perioperatively vaccinated and comparator patients. Surgical treatment was also included in the propensity-matching model, along with tumor anatomy, MGMT/IDH status, frailty, ASA, KPS, socioeconomic deprivation, and surgery year. After matching, baseline covariates were well balanced and the survival difference was still observed.

Additional surgical sensitivity analyses further supported this interpretation. Models incorporating GTR and non-GTR indicators, as well as resection-only models, yielded consistent results after extent-of-resection and treatment-proxy adjustment. These analyses do not replace volumetric residual-enhancement assessment, which was not uniformly available, but they directly address the most obvious surgical confounders: biopsy versus resection and extent-of-resection category.

### Negative-control vaccination

Healthy-vaccinee bias remains an important limitation of any observational vaccination study. Vaccinated patients may be more health-engaged, more connected to care, less frail, more adherent to therapy, or more likely to complete adjuvant treatment. We addressed this concern using measured clinical adjustment, socioeconomic variables, treatment-completion analyses, and negative-control vaccination analyses.

Influenza vaccination within one year before surgery was not associated with improved survival. In an analogous 100-day preoperative window, influenza vaccination again was not associated with improved survival. Additional non-COVID vaccine proxies also did not reproduce the COVID-vaccination association. Pneumococcal vaccination and any documented non-COVID vaccination were null, and zoster vaccination was associated with worse rather than improved survival. These negative-control analyses do not prove causality, but they make the finding harder to dismiss as a generalized “vaccinated patients live longer” effect.

### Immune status, steroids, and COVID severity

Another possible explanation is that perioperatively vaccinated patients had better baseline immune reserve, lower steroid exposure, or less severe COVID-related illness. This is especially important in GBM, where lymphopenia, myeloid predominance, and corticosteroid exposure may influence both treatment tolerance and antitumor immunity.^27,28^

The association remained after adjustment for perioperative steroid exposure, absolute lymphocyte count, and COVID hospitalization/severity. It also remained present after excluding patients with COVID hospitalization. These findings suggest that the observed survival association is not explained solely by severe COVID avoidance, steroid exposure, or baseline lymphocyte differences. Nevertheless, EHR-derived steroid and CBC variables remain imperfect proxies for dynamic immune function and cumulative immunosuppressive burden. *Surgery and antigen release*

Initial surgical management of GBM is usually understood in terms of diagnosis, cytoreduction, decompression, and treatment planning. However, the perioperative period is also a distinct immunologic event. Tumor tissue is disrupted, tumor antigens are released, local inflammation and wound-healing programs are activated, the blood-brain barrier and tumor-brain interface are perturbed, corticosteroids are frequently administered, and lymphocyte dynamics shift. This makes first tumor surgery a plausible biologic window in which systemic immune tone could matter.^9^

The perioperative window is especially relevant in GBM because the tumor microenvironment is immunologically hostile. GBM is enriched for tumor-associated macrophages and microglia, impaired antigen presentation, T-cell dysfunction, immune exclusion, and steroid-sensitive lymphopenia. These factors have made GBM a poor responder to checkpoint blockade and many tumor-directed vaccine approaches.^5–8,29^ A recent systemic immune stimulus before surgery may not reverse this immunosuppression, but it could alter the immune context in which tumor antigen release occurs.

This is the key biologic premise of the study: a non-tumor-specific vaccine may matter not because it targets GBM directly, but because it changes the immune state of the patient at the moment tumor antigens become available.

### mRNA vaccination as systemic immune priming

The perioperatively vaccinated cohort was overwhelmingly mRNA-vaccinated. Adenoviral-vector recipients had median overall survival of 639 days, which did not suggest an obviously adverse platform-specific pattern. However, because only 5 perioperatively vaccinated patients received adenoviral-vector vaccines, this study should be interpreted as an mRNA-dominant exposure analysis rather than a comparison of vaccine platforms. This is relevant because the mechanistic hypothesis is not limited to prevention of SARS-CoV-2 infection. mRNA/lipid nanoparticle vaccines can activate innate immune pathways, type I interferon signaling, dendritic-cell activation, and downstream T-cell priming.^10,11^ Recent work in non-CNS malignancies has shown that SARS-CoV-2 mRNA vaccination can sensitize tumors to immune checkpoint blockade, with preclinical data implicating type I interferon–mediated innate immune activation and enhanced CD8-positive T-cell priming against tumor-associated antigens.^12^

Our findings are consistent with that broader immunologic framework, but they do not prove it. The vaccine does not need to encode a GBM antigen to be relevant. Instead, recent mRNA vaccination may be associated with a systemic immune state that intersects with perioperative tumor antigen release. In this model, the potential interaction is between vaccine-associated immune activation and surgery-associated antigen availability, rather than between SARS-CoV-2 spike protein and a specific GBM antigen.

### Exploratory shared-epitope findings

In addition to nonspecific systemic immune priming, the exploratory computational analyses identified a specific HLA-A*02:01-restricted shared-epitope candidate between SARS-CoV-2 spike and the GBM-associated antigen NLGN4X. The NLGN4X peptide NLDTLMTYV and spike peptide RLQSLQTYV shared 5 of 9 amino acids, including both HLA-A*02:01 anchor positions and an identical C-terminal T-Y-V motif. Both peptides were strong predicted HLA-A*02:01 ligands. NLGN4X NLDTLMTYV has previously been identified in the HLA-A*02:01-associated GBM peptidome, and NLGN4X-directed TCR-transgenic T cells have demonstrated recognition of peptide-loaded and naturally NLGN4X-expressing glioma models.^30,31^ Thus, NLGN4X represents a biologically established GBM antigen rather than a source protein selected solely through sequence matching.

Public TCGA and CGGA data further showed that NLGN4X was expressed across independent GBM cohorts. Expression alone, however, does not establish intracellular processing or cell-surface presentation of NLDTLMTYV. NLGN4X-high tumors also were not consistently immune-inflamed in bulk transcriptomic analyses and did not have reproducibly different survival. These observations suggest that the potential relevance of NLGN4X lies in its role as an available HLA-restricted antigen rather than as a stand-alone prognostic or immune-infiltration marker.

The structural analyses extended the candidate beyond linear sequence similarity. The modeled NLGN4X and spike peptides adopted closely overlapping HLA-A*02:01-bound backbones and similar placement of the shared P5, P7, and P8 side chains. NLGN4X also remained geometrically compatible when introduced into the experimentally determined RLQ-specific TCR/HLA-A2 footprint.^39^ The substitution at P6 from glutamine in spike to methionine in NLGN4X remains an important uncertainty because P6 participates in the experimental TCR interface. The tested influenza HA comparator peptides did not reproduce the same combined profile of strong same-HLA binding, close peptide-HLA structural concordance, and compatibility with the RLQ-specific TCR footprint.

To our knowledge, this is the first report to nominate the NLGN4X-derived peptide NLDTLMTYV and SARS-CoV-2 spike-derived peptide RLQSLQTYV as a candidate HLA-A*02:01-restricted shared epitope in GBM. This observation provides a concrete and experimentally testable molecular-mimicry hypothesis, but it should not be interpreted as evidence that this mechanism caused the clinical survival association. The computational screen was post hoc, candidate-based, and limited to predicted and modeled molecular properties. Functional cross-reactivity requires demonstration that the same HLA-A*02:01-restricted T-cell population can recognize both peptides.

### Early postoperative course

The perioperative utilization findings were secondary but directionally consistent with the survival analysis. Perioperatively vaccinated patients had shorter median length of stay and numerically lower 30-day readmission, although the readmission difference did not reach statistical significance. The LOS difference was small in absolute terms and was not explained by a higher biopsy rate in the vaccinated group, as biopsy-versus-resection distribution was nearly identical between groups. Instead, the LOS distribution suggested fewer prolonged postoperative stays among vaccinated patients.

These findings should not be interpreted as evidence that vaccination reduced postoperative complications. Length of stay is affected by tumor anatomy, neurologic status, discharge disposition, rehabilitation placement, surgeon practice, social support, and postoperative complications. Still, the utilization findings support the narrower conclusion that perioperative vaccination was not associated with a worse early postoperative course.

### Clinical implications

These findings should not be interpreted to mean that urgent GBM surgery should be delayed solely to administer a COVID-19 vaccine. Timely surgery remains essential for diagnosis, decompression, cytoreduction when feasible, and initiation of standard adjuvant therapy. The practical implication is narrower and more defensible: vaccination status may be a relevant component of preoperative optimization when timing allows.

If validated, this study suggests that being recently vaccinated before first GBM surgery may have relevance beyond prevention of severe COVID-19. Because vaccination status is modifiable, routinely documented, and already part of general medical care, it could become a low-burden perioperative counseling point. However, any formal recommendation would require prospective validation and should not supersede standard neurosurgical urgency.

### Relationship to GBM immunotherapy

Therapeutic GBM vaccines and checkpoint inhibitors have produced inconsistent clinical benefit, largely because of tumor heterogeneity, immune exclusion, steroid exposure, myeloid dominance, and systemic immune dysfunction.^7,8,29^ The present study does not contradict that experience. Instead, it raises a different question: whether immune timing matters.

Tumor-specific vaccines attempt to create targeted antitumor immunity.^8,29^ Perioperative COVID-19 vaccination, in contrast, may mark a nonspecific immune-activated state. The potential relevance lies in the coincidence of that state with surgery. If tumor antigen release occurs while innate and adaptive immune pathways are recently activated, even a non-tumor-specific vaccine could plausibly influence antitumor immune surveillance. This framing may help explain why a routine public-health intervention could show an association with GBM survival despite not being designed as a cancer therapy.

### Mechanistic validation and planned functional testing

The present study remains clinical and observational, and the computational analyses do not prove the mechanism. Tissue-based immune profiling remains an important near-term follow-up. Archival tumor tissue from perioperatively vaccinated and comparator patients could be assessed for CD8-positive T-cell infiltration, macrophage/microglial phenotype, MHC-I expression, PD-L1 expression, granzyme B, and interferon-response markers such as MX1 or ISG15.^5,12^ HLA typing and tumor immunopeptidomics would be particularly informative because they could determine whether NLDTLMTYV is naturally presented by HLA-A*02:01-positive GBM specimens.

The most direct functional test of the shared-epitope hypothesis would be a staged HLA-A*02:01-restricted peptide cross-reactivity assay. Peripheral blood mononuclear cells from HLA-A*02:01-positive, COVID-vaccinated donors would be stimulated or expanded with spike RLQSLQTYV and then challenged separately with RLQSLQTYV, NLGN4X NLDTLMTYV, the influenza comparator peptides, and an irrelevant HLA-A*02:01-binding peptide. Initial readouts would include IFN-γ ELISpot and flow-cytometric assessment of CD8-positive T-cell activation, including CD69 or CD137 induction, intracellular IFN-γ/TNF-α, and CD107a degranulation. An anti-HLA-A2 blocking condition would test whether any shared response was HLA-A*02:01 dependent.

If the initial assay identifies reproducible dual-peptide reactivity, confirmatory studies would use HLA-A*02:01/RLQSLQTYV and HLA-A*02:01/NLDTLMTYV peptide-MHC tetramers or dextramers to isolate antigen-reactive cells.^40^ T-cell receptor sequencing could then determine whether overlapping clonotypes recognize both peptide-HLA complexes. A final tumor-relevance experiment would test whether spike-reactive T cells recognize or kill HLA-A*02:01-positive, NLGN4X-expressing GBM cells, with HLA-A2 blockade, NLGN4X knockdown, HLA-A*02:01-negative tumor cells, and peptide-pulsed antigen-presenting cells as controls. This staged design would distinguish peptide-level cross-reactivity from endogenous tumor-cell recognition.

### Limitations

This study has several limitations. First, it is retrospective and single-center, and causality cannot be inferred. Although propensity matching, negative-control analyses, treatment-completion sensitivity analyses, calendar-year analyses, and secondary covariate adjustment reduce measured confounding, residual confounding remains possible. Second, vaccination ascertainment required integration of structured immunization records with manual review of outside primary-care and pharmacy documentation. This improves exposure classification but may introduce heterogeneity in documentation completeness. Third, KPS and ECOG were extracted from clinical notes and clinical event records, and performance-status documentation was not uniformly timed in all patients. Timing was recorded and sensitivity analyses were performed but note-derived functional status remains imperfect.

Fourth, steroid exposure and perioperative CBC variables were derived from available EHR data within a perioperative window and may not fully capture cumulative immunosuppressive exposure or dynamic immune function.^27,28^ Fifth, although the exposure was predominantly mRNA-based, the study was not powered to compare vaccine platforms directly. The platform-specific interpretation should therefore remain cautious. Sixth, adjuvant treatment variables were evaluated using available treatment proxies. Radiotherapy receipt, temozolomide exposure, TTFields/Optune use, and a combined Stupp-regimen proxy were incorporated into sensitivity analyses, but exact temozolomide cycle counts, radiotherapy interruptions, TTFields adherence, and salvage therapy sequencing were not uniformly available. Therefore, treatment completion and dose intensity cannot be fully disentangled from the observed survival association.

Seventh, tumor volume, eloquent-cortex involvement, and volumetric residual-enhancement measures were not uniformly available. We addressed surgical-selection bias using biopsy versus resection, extent-of-resection category, tumor location, multifocality, and deep/midline involvement, but residual anatomic confounding may remain. Finally, the study does not include patient-level HLA typing, direct tumor immunopeptidomics, T-cell receptor sequencing, or functional T-cell cross-reactivity assays. The public transcriptomic and structural analyses were post hoc and hypothesis-generating; they used bulk expression datasets, a curated peptide panel, predicted HLA binding, and representative peptide-HLA models without molecular-dynamics simulation or binding free-energy estimation. These analyses support candidate prioritization but do not establish endogenous peptide presentation, shared TCR recognition, or a causal mechanism for the clinical survival association.

### Future validation

The next step is multicenter validation. A multi-institutional cohort would test whether the association persists across different vaccine-capture systems, neurosurgical practices, neuro-oncology treatment patterns, patient populations, and COVID-era calendar effects. Larger numbers would also allow better evaluation of mRNA-only exposure, booster status, steroid-stratified analyses, resection-only cohorts, treatment-completion endpoints, and timing thresholds.

Prospective validation should be paired with tissue and blood-based immune studies. The highest-yield translational design would compare perioperatively vaccinated and matched unvaccinated GBM patients using tumor immunohistochemistry, multiplex immunofluorescence, RNA-expression signatures, and peripheral immune profiling. This would directly test whether the clinical association corresponds to measurable immune activation at the tumor or systemic level.

## Conclusion

Perioperative COVID-19 vaccination within 100 days before first tumor surgery was associated with improved overall survival in newly diagnosed glioblastoma. The association persisted after propensity matching and across sensitivity analyses addressing adjuvant treatment, calendar time, surgical selection, perioperative steroid exposure, immune-cell variables, COVID severity, and negative-control vaccination. These findings suggest that immune activation timing relative to GBM surgery may be clinically relevant and identify perioperative vaccination as a previously underexplored variable in GBM outcomes. Multicenter validation and tissue-based immune profiling are warranted.

## Data availability

Deidentified clinical data and the associated computational materials supporting the findings of this study are available from the corresponding author upon reasonable request, subject to University of Alabama at Birmingham approval, any required data-use agreement, and applicable patient-privacy, ethical, and third-party data-use restrictions.

